# East-West mortality disparities during the COVID-19 pandemic widen the historical longevity divide in Europe: an international comparative study

**DOI:** 10.1101/2023.11.08.23298275

**Authors:** Vladimir M. Shkolnikov, Sergey Timonin, Dmitry Jdanov, Naomi Medina-Jaudes, Nazrul Islam, David A. Leon

## Abstract

**Introduction:** For over half a century, life expectancy in the former communist countries of Eastern Europe has been noticeably lower than in Western Europe. Since 2000 this gap has narrowed. We examined the impact of the COVID-19 pandemic on these long-term mortality trends and differences.

**Methods:** Nationally reported mortality data were used to estimate East-West differences in excess mortality and life expectancy losses. Regression and decomposition methods were employed to examine the contribution of vaccination, trust in government, regulatory enforcement, and air connectivity between populations to these differences.

**Results:** During the pandemic, the East-West life expectancy gap widened to its highest level in more than two decades. Moreover, the trajectory of excess mortality during the pandemic differed between East and West, with levels of excess mortality in the East being minimal until autumn 2020. Cumulative excess mortality in weeks 10-18 of 2020 was correlated with an index of air connectivity, which was appreciably lower in Eastern compared to Western European countries in the immediate pre-pandemic period. From October 2020 onwards, the East suffered greater losses in life expectancy, especially in 2021. This could not be explained by greater frailty of the Eastern European populations, as indicated by higher pre-pandemic mortality levels. Half of the difference between East and West in 2021 was jointly explained by COVID-19 vaccination levels and trust in government.

**Conclusions:** East-West contrasts in the timing and magnitude of life expectancy losses during the COVID-19 pandemic appear to have their ultimate origins in differences between societies that were established during the Cold War. These include differences in the connectivity of populations, levels of trust in science and authorities and the related capacity of Eastern countries to mount effective vaccination campaigns.

## INTRODUCTION

Since the late 1960s, life expectancy in the former communist countries of Eastern Europe has lagged behind those of Western Europe, although from 2000 this gap started to narrow.^1^ As previously reported, the overall levels of pandemic excess mortality in Eastern European countries in 2020-21 were higher than in Western countries.^2–10^ Schöley and colleagues noted that in Europe, the East-West differences in life expectancy losses were larger in 2021 than in 2020.^10^ They importantly also found that life expectancy losses in the fourth quarter of 2021 were correlated with vaccination coverage.

In addition to establishing differences in overall excess mortality between East and West, some studies have noted that compared to Western European countries those of the East had a relatively mild first phase of the pandemic, with excess deaths only climbing steeply from autumn 2020 onwards.^11^ In contrast, many Western countries were hit hard in the spring of 2020 but managed to avoid a large increase in the autumn/winter of 2020 and early 2021.^9^ ^10^ This difference in the timing of major impacts of the pandemic between East and West has not been systematically investigated.

Apart from differences in vaccination coverage between the East and West, which would only have impacted the trajectory of the pandemic from 2021 onwards, several other factors have been mentioned as explaining East-West differences in pandemic mortality. Inadequate or ineffective non-pharmaceutical interventions (NPIs), as well as low trust in government and science have been considered as central.^12^ Individual-level evidence from Germany linked low trust among older individuals with exposure to communism.^13^ The lower ability of Eastern European authorities to enforce regulatory measures may also have contributed to higher life expectancy losses in the East.^14^

We have examined life expectancy losses and excess mortality due to the COVID-19 pandemic by comparing the higher losses in the group of 11 Eastern European countries of the former communist bloc with the lower losses in 17 Western European countries. We particularly examined the association between air travel connectivity between European countries and the timing of the pandemic’s impact in Eastern compared to Western Europe. We also analysed the association between population-level factors and East-West contrast in life expectancy losses in 2021. These include vaccination levels, the stringency of NPIs, trust in government/science, and the effectiveness of regulatory enforcement. We also quantified the contribution of baseline mortality and excess mortality to the observed difference in life expectancy between the East and West. Finally, we have reflected on how much any of these potential explanatory factors may have their roots in the very different post-war histories of Eastern and Western Europe.

## METHODS

We used annual mortality data for European countries up to and including 2021 from the Human Mortality Database (HMD)^15^ and weekly mortality data from the Short-Term Mortality Fluctuations (STMF) series.^16^ We divided the resulting 28 populations into two groups: Eastern Europe (East) consisting of 11 former communist countries and Western Europe (West) consisting of the remaining 17 countries (Supplementary Table S1). To analyse the temporal pattern of excess mortality, we identified for each country the month of the first major peak of the excess death rate (EDR) and the total EDR over 2020-21 (see Supplementary Data and Methods), using maps to visualize these trends.

Commonly used approaches to calculating longevity differences between two periods (e.g. 2019 and 2020-21) can underestimate the scale of true losses. ^9,10^ To avoid this, we compared life expectancies observed in 2020-21 with the expected values forecasted from past mortality trends that would have been observed in the absence of the pandemic (Supplementary Figure S1). We used the Lee-Carter model to estimate the expected mortality using the period 2005-19 as the baseline, having previously undertaken sensitivity analyses with respect to the choice of baseline period.^8, 11^ We calculated the life expectancy losses as the differences between the observed and expected life expectancies in 2020 and 2021, overall, and also split into two broad age groups: 0-64 years and 65+ years (see Supplementary Data and Methods).

We quantified the contributions of the baseline mortality (pre-existing population health) and the relative (proportional) mortality excess (the pandemic *per se*) to the change in the East-West difference in life expectancy losses. A counterfactual method was used to calculate the corresponding *Level* and *Change* components (see Supplementary Data and Methods).

Whether a country had a major excess mortality peak in March-April 2020 reflects the number of cases in each country immediately prior to the introduction of wide ranging measures to reduce transmission within their population. If the number was low, then the sharp reduction in person-to-person mixing that resulted from lock-down measures might have been sufficient to prevent the establishment of a self-sustaining, exponential growth in cases. Within Europe, by February 2020, most of these initial cases would have been imported from other European countries, such as Italy which experienced the earliest peak in the region, as direct flights from China to Europe had already been severely reduced. We therefore examined the likelihood of the introduction of cases into each country from other European countries in the immediate pre-pandemic period using an index of air passenger connectivity. We hypothesise that the number of such imported cases and the subsequent excess mortality may be related to the travel connectivity of each country to others in the region. We assessed rank order correlations for European countries between EDRs in March-April (weeks 10 to 18) of 2020 and the average daily numbers of arriving flights from other European countries in the period February 10-23, 2020 using data from the OpenSky network^17^ (see Supplementary Data and Methods).

We examined the potential contribution of population-level factors to explaining the very substantial East-West contrast in life expectancy losses in 2021. These factors were the stringency index, vaccination coverage, trust in government, trust in science, and a regulatory enforcement score (see Supplementary Data and Methods).^18–21^ Our analysis was guided by a simple conceptual framework (Figure 1) relating various factors to 2021 losses. Proxy data for a number of these factors (marked in green) were used in a meta-regression framework. We regressed sex-specific life expectancy losses in 2021 to see how this difference was attenuated in response to adjustment for single explanatory variables and for two-variable combinations. As described in the Supplementary Methods, we checked the normality of the distributions and performed statistical tests for linearity and heteroscedasticity for each explanatory variable.

**Figure 1.**
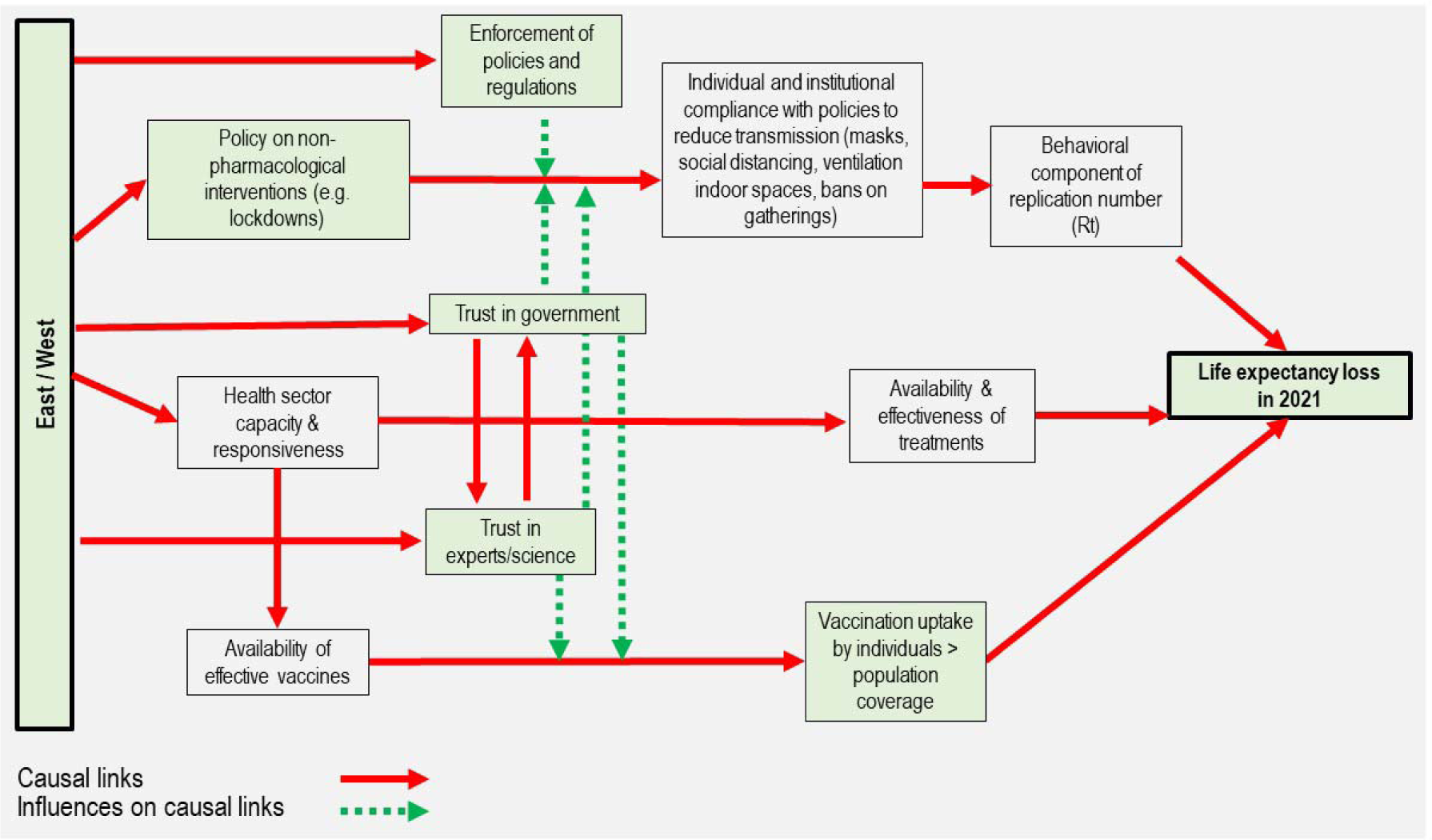
Conceptual diagram: factors that may explain the East-West gap in life expectancy losses in 2021. The green color of the boxes indicates the availability of numerical proxy variables.

Life table calculations were carried out in R (version 4.2.2); the rest of the statistical analysis was conducted in Stata 17. The scripts and corresponding data and calculations are available at: https://github.com/VMSdemo/East-West-contrast-in-life-expectancy-losses-in-2020-21. Maps were built in ArcGIS 10.8.2.

## RESULTS

### Life expectancy trends since 2000 and abrupt changes in 2020-21

The long-term life expectancy disadvantage of the East compared to the West began to reduce in the late 1990s in the new EU member states of Central and Eastern Europe, and in the mid-2000s in Russia (Supplementary Figure S2). Between 2000 and 2019, the difference in average life expectancies between the East and West decreased from 7.7 years to 6.3 years for males, and from 4.4 years to 3.2 years for females. This convergence was reversed by the COVID-19 pandemic: in 2020, the East-West difference increased to 6.5 years for males and 3.5 years for females. In 2021, the gap widened further to 7.9 and 4.9 years, respectively. This was caused by a slightly larger fall in life expectancy in the East than in the West in 2020 (males: 0.9 years Eastern Europe vs 0.7 Western Europe; females: 0.8 years and 0.5 years, respectively). In 2021, the East saw a continuing fall of 1.3 years for both males and females, while the West experienced a small rebound with a gain of 0.1 years in both sexes. All Eastern European countries had larger declines in 2021 than in 2020.

### Temporal and geographic patterns of excess mortality

The weekly EDRs in 2020-21 by country are shown in Figure 2 together with the Eastern and Western mean EDRs. The countries with the highest EDR peaks in the first pandemic wave (March-May 2020) were in Western Europe. During this initial phase, the pandemic had little impact on mortality in countries of the East, except Russia, which experienced moderate excess mortality in May-June 2020. This changed in October 2020 when most Eastern countries experienced marked increases in weekly excess deaths. From November 2020 to January 2021, the EDR peak was higher and wider in the East than in the West. Most importantly, countries of the East experienced further massive elevations of weekly EDRs throughout 2021.

**Figure 2.**
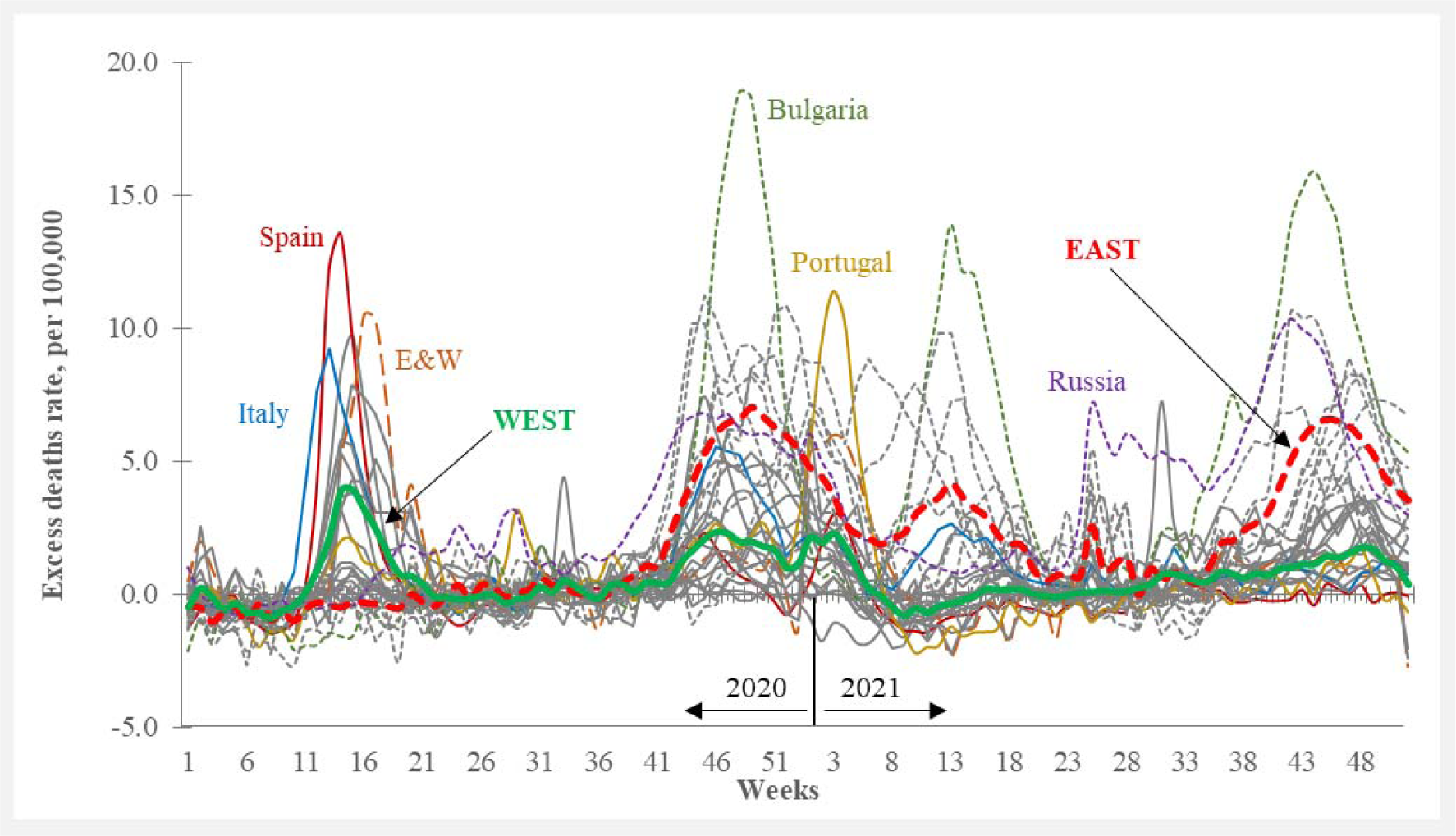
Weekly excess death rates in 2020 and 2021, total population. Western European countries showed the highest mortality peaks in the first pandemic wave (March-May 2020), while Eastern European countries showed marked increases in weekly excess deaths only in October 2020 - January 2021 and further experienced large elevations of excess mortality throughout 2021. Data shown in this Figure is provided at https://github.com/VMSdemo/East-West-contrast-in-life-expectancy-losses-in-2020-21

Panel A of Figure 3 shows that the first major peak in EDR in each country moved over time from the southwest to the northeast of Europe. The earliest major excess mortality waves in the spring of 2020 occurred in Italy, Spain, England and Wales, Scotland, Northern Ireland, Belgium, the Netherlands, France, and Sweden. Most other Western countries and nearly all Eastern ones experienced later major peaks between November 2020 and January 2021.

**Figure 3.**
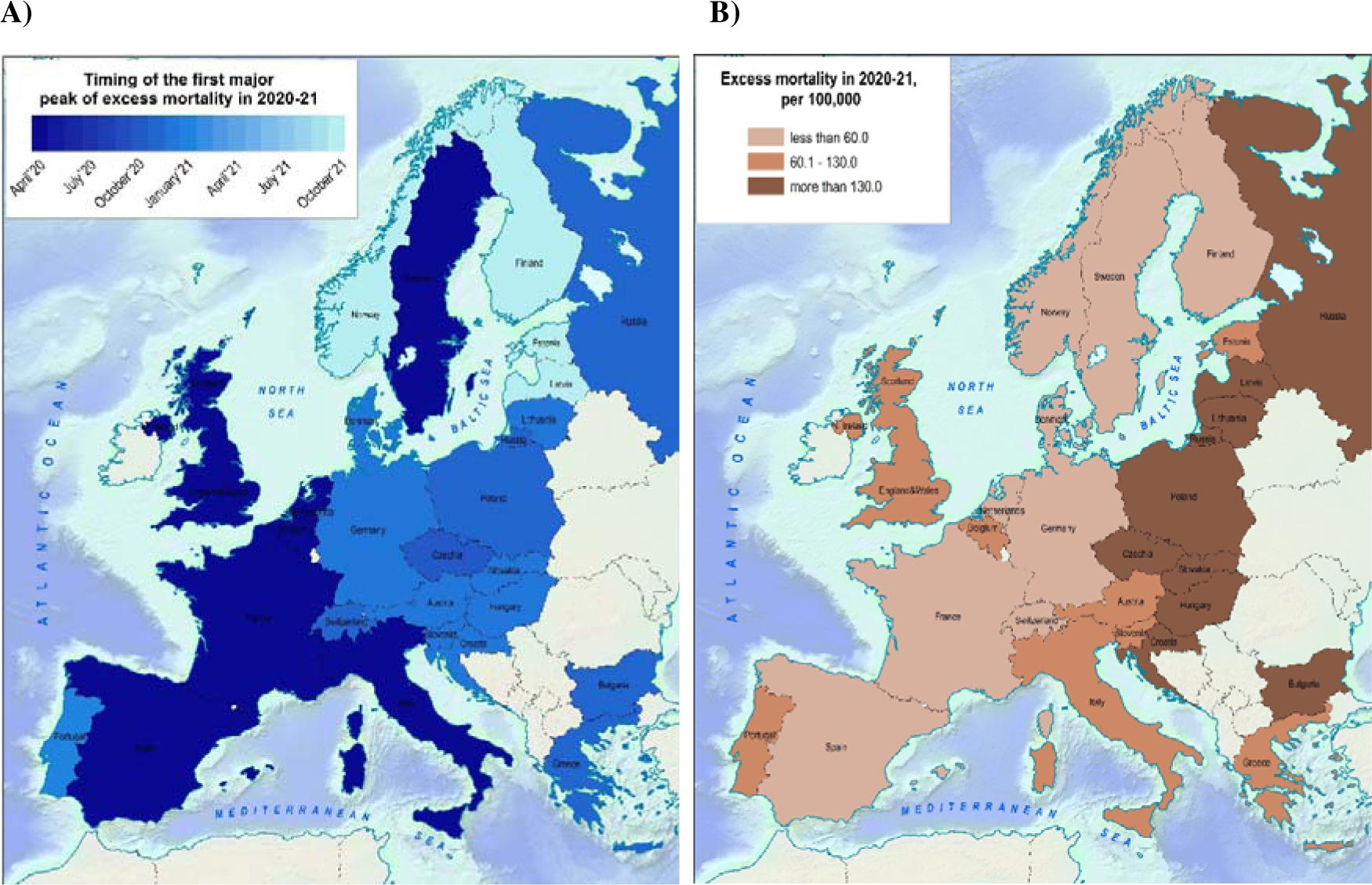
Spatial pattern of excess mortality during the COVID-19 pandemic. A) Timing of the first major peak. B) Excess death rates in 2020-21. In general, the first major excess mortality peak moves from the southwest to the northeast of Europe. Despite the later onset of excess mortality in Eastern compared to many Western European countries, the East suffered much higher total excess death rates in 2020-21. Data shown in this Figure is provided at https://github.com/VMSdemo/East-West-contrast-in-life-expectancy-losses-in-2020-21

Despite the earlier onset of excess mortality and its higher level in spring 2020 in many Western countries, total mortality excess in 2020-21 was substantially higher in Eastern countries than in Western ones, except for Slovenia (Figure 3, Panel B). It is particularly striking that while the Western countries situated in the center of Europe, such as Germany, Austria, and Switzerland, had similar timings in their first pandemic peaks as neighboring countries in the East (Poland, Czechia, Hungary, and others), their overall rate of excess mortality in 2020-21 was much lower. Even Latvia and Estonia, which had very late excess mortality onset in October-November 2021, similar to trends in Norway and Finland, experienced much higher total excess mortality in 2020-21 than any Western country.

Supplementary Table S2. demonstrates the higher intensity of arriving flights in Western compared to Eastern European countries. In the immediate pre-pandemic period (10-23 February 2020), the mean numbers of flights per day arriving in all Western European countries from another European country was 6356, compared to 816 in all Eastern countries. Supplementary Figure S3 suggests a positive relationship between the excess death rate in March-April 2020 and the flight connectivity in mid-February 2020: Spearman rho of 0.782 (p<0.001) (95%CI 0.627,0.936) (more in the Supplementary Methods). The main outlier is Germany, which is exceptional as Frankfurt is by far its busiest airport and is one of the largest transit hubs in the world.

### Life expectancy losses in 2020-21 and the reemergence of the East-West divide

Life expectancy losses in 2020 overlapped between East and West (Supplementary Table S3). However, in 2021, the losses were substantially higher in the East (2.89 95%CI (2.65,3.13) years for males and 2.52 95%CI (2.30,2.74) years for females) than in the West (0.97 95%CI (0.80,1.16) years for males and 0.66 95%CI (0.47,0.86) years for females) (see also Supplementary Figure S4). Notably, the mean life expectancy losses in Western Europe in 2021 were below the minimum country-specific losses in Eastern countries in both sexes (Supplementary Table S3).

While there was already a tendency for higher life expectancy losses in Eastern countries in 2020, this became more pronounced in 2021. The upper panels of Figure 4 show that countries with the largest life expectancy losses in 2020 included both Western (Italy, Spain, parts of the UK, and Belgium) and Eastern (Russia, Lithuania, Bulgaria, Poland, Czechia) countries. However, the lower panels of Figure 4 demonstrate that in 2021, the ten countries with the highest life expectancy losses were all from Eastern Europe.

**Figure 4.**
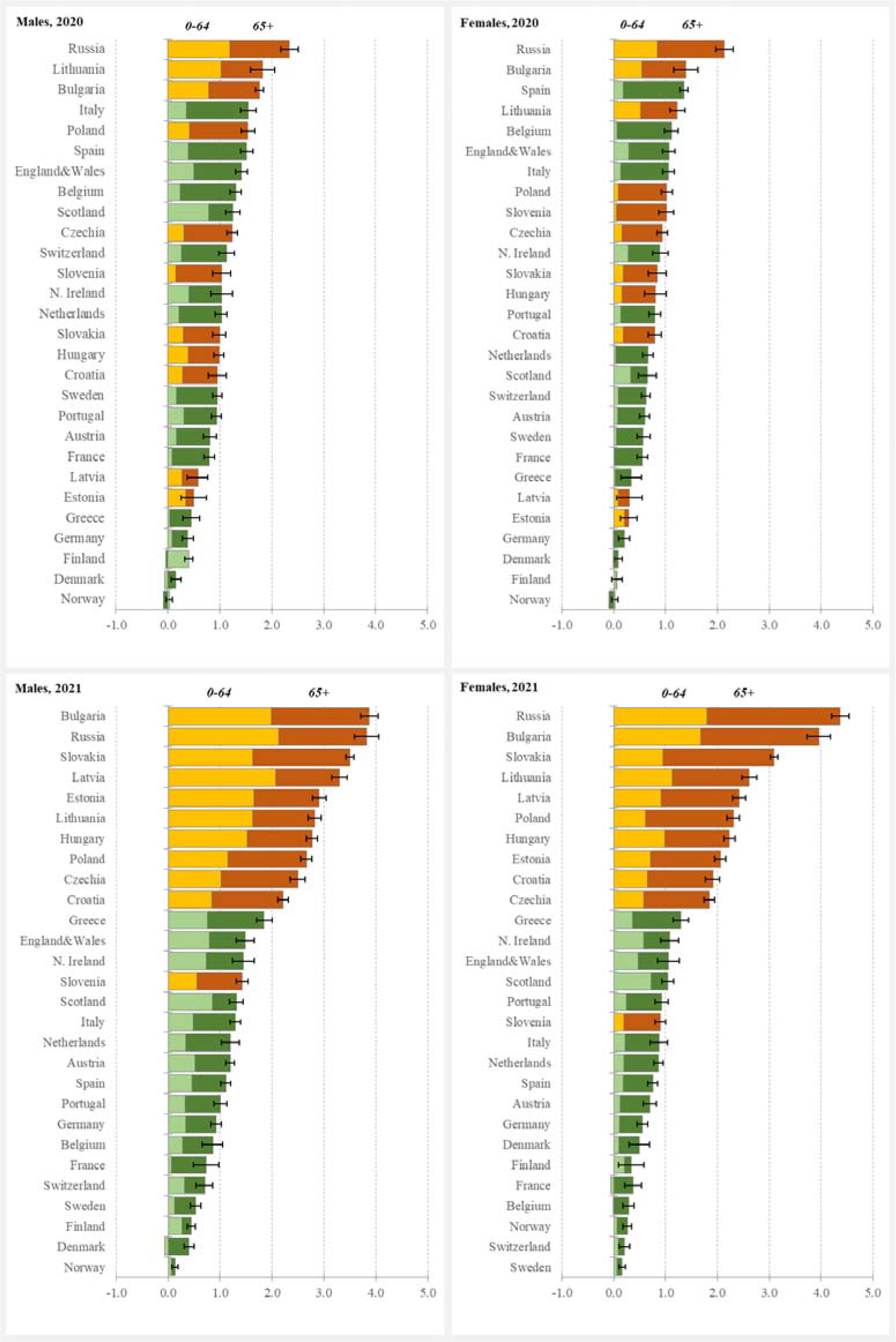
Life expectancy losses and their age components, by year and sex, in years. The figure shows the ranking of Eastern (yellow-brown colour) and Western (green) European countries by life expectancy losses in 2020-21 as well as the contribution of mortality changes below and above age 65 to the loss. The contribution of younger ages was generally higher in males, in 2021 and in the East. Data shown in this Figure is provided at https://github.com/VMSdemo/East-West-contrast-in-life-expectancy-losses-in-2020-21

Figure 4 also shows the contribution of different age groups to overall losses for each country. Overall, in 2020, the contributions of excess deaths occurring under the age of 65 years to total life expectancy losses were 31.5% for males (25.3% in the West vs. 39.5% in the East) and 23.2% for females (19.3% in the West vs. 28.5% in the East). In 2021, deaths under 65 years made up 44.1% of the total losses of life expectancy for males (40.3% in the West vs. 49.7% in the East) and 35% for females (34.8% in the West vs. 35.2% in the East).

### Can initial differences in population health explain East-West differences in 2021?

The East-West differences in the impact of the COVID-19 pandemic on excess mortality and life expectancy losses can be separated into those resulting from larger proportional increases in mortality in the East compared to the West and those due to the higher baseline mortality in the East reflecting the poorer health status of Eastern European populations.

The average difference in the life expectancy losses in 2021 between the East and the West was 2.03 years for males and 1.97 years for females. As shown in Supplementary Table S4 the contributions of larger relative mortality excess in the East to these differences were 88% for males and 94% for females. Further supporting evidence for a substantial East-West difference in relative increases in mortality is provided in Supplementary Figure S5. These analyses lead us to conclude that it is the larger relative increases in mortality that is the primary driver of the life expectancy losses, rather than higher levels of baseline mortality in the East compared to the West.

### Associations of explanatory factors with East-West differences in life expectancy losses in 2021

The scatterplots in Supplementary Figure S6 show statistically significant correlations between life expectancy losses in 2021 with country-level factors (explanatory variables), except for the stringency index. Table 1 shows the effect of adjustment for a series of factors on the East-West difference in life expectancy losses in 2021. Without adjustment for any factors, the losses of life expectancy were about 2 years greater in Eastern compared to Western Europe. Vaccination coverage on 1 September 2021 led to the largest attenuation of this effect of any single variable. Regulatory enforcement and trust in government had the second and third largest impacts on the East-West differences, respectively. The stringency index of NPIs did not attenuate the association at all. Vaccination in combination with trust in government showed the largest reduction of any model, resulting in a halving of the East-West differences in life expectancy losses. Adjustment for vaccination together with regulatory enforcement had a similar effect for females, but a slightly smaller attenuating effect for males. The two-factor models had a marginally better fit than the single-factor (vaccination) models.

**Table 1.**
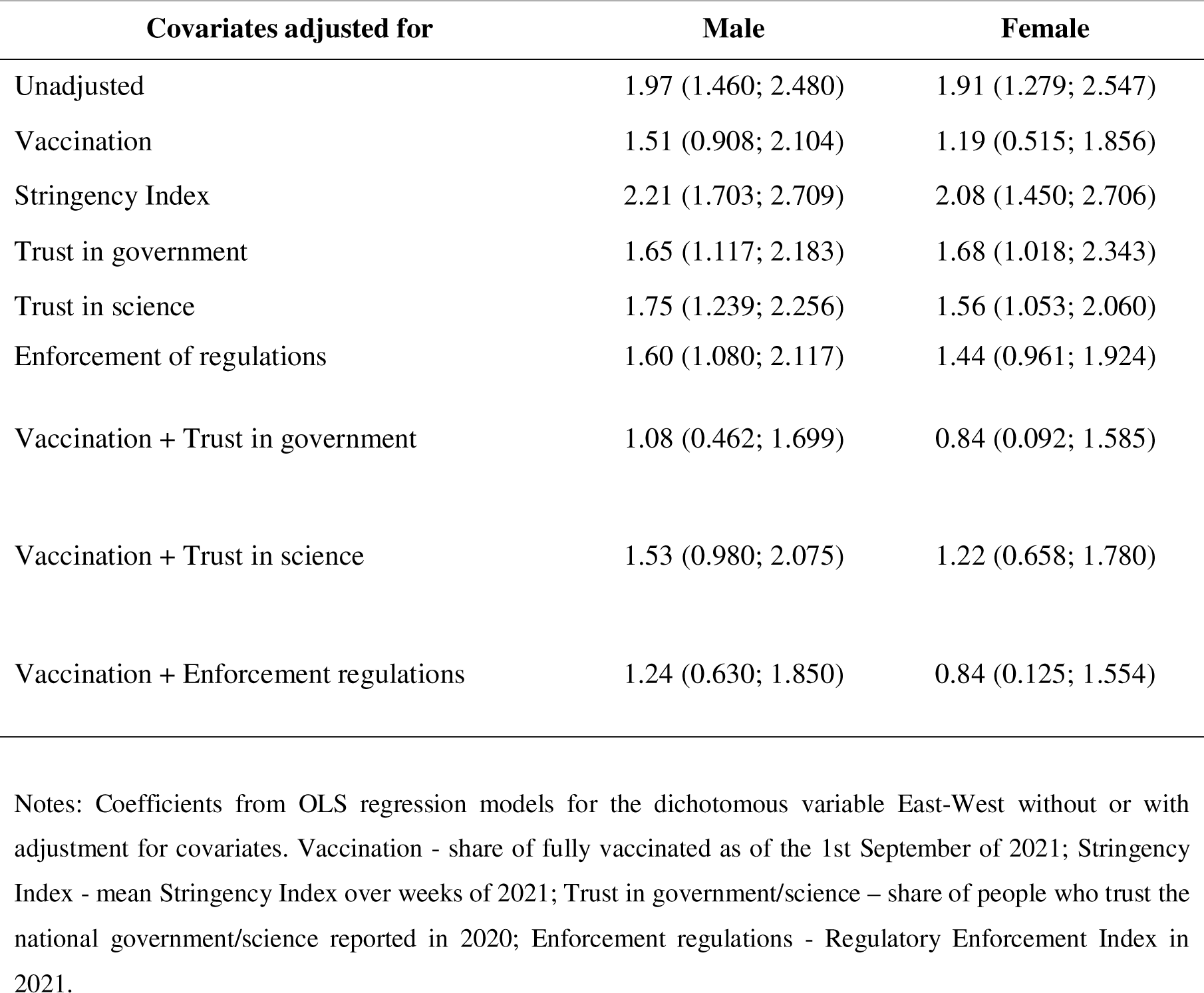
The model-based East-West difference in life expectancy losses in 2021, in years.

## DISCUSSION

We have found that the differential impacts of the COVID-19 pandemic on excess mortality and life expectancy losses in Eastern compared to Western Europe have two distinctive features. First is the absence in nearly all Eastern European countries of a pronounced excess mortality in the first few months of the pandemic unlike in many Western European countries. Second, from October 2020 to the end of 2021, most Eastern European countries tended to experience more persistent high levels of excess mortality compared to Western European countries. This more than offset the approximately 6-month-later onset of substantial excess deaths in Eastern European countries, resulting in a much greater overall toll of excess deaths and life expectancy losses than in Western Europe in the period up to the end of 2021.

The initial low levels of excess mortality in Eastern Europe seen in the first months of the pandemic when many Western European countries, such as Italy, Spain, and the UK, had massive excess mortality peaks has been noted by others.^9–11^ However, little attention has been given to explaining why this might be the case. Countries with low levels of excess mortality in March and April 2020 are likely to be those that had a low prevalence of COVID-19 at the time when they imposed various degrees of restriction and lockdown in March 2020. On the other hand, those with a high prevalence at this point would be more likely to be subject to exponential spread of the virus. Direct estimates of the prevalence of COVID-19 infection have been challenging throughout the pandemic. Several attempts have been made to estimate the prevalence of infection per capita in the early stages of the pandemic. Russell et al.^16^ used mortality from COVID-19 to work backwards to estimate the prevalence of infection several weeks earlier, making a series of assumptions about case-fatality and the period from infection to death. In mid-March 2020, when nearly all countries had introduced their first range of public health measures/lockdowns, the estimated prevalence of COVID-19 infection was systematically lower in Eastern compared to Western countries.

As all initial cases in a country of this novel virus will have been the result of importation, the degree of cross-border travel in the immediate pre-pandemic period will be an important driver of early phase prevalence. One index of connectivity is the number of inbound passenger flights. We have found that there is a positive correlation across countries between the immediate pre-pandemic passenger flight connectivity between European countries and excess mortality in March-April 2020, with the level of flight connectivity and excess mortality being far lower for Eastern compared to Western European countries. The low level of connectivity in the East could have led to a much lower prevalence of COVID-19 cases in March 2020 than most countries. We interpret this as supporting the hypothesis that in the weeks leading up to the imposition of various degrees of lockdown in March 2020, the scale of the epidemic spread of COVID-19 in each country would be proportional to the number of prevalent cases in the population. If this were low, the imposition of steps to reduce person-to-person transmission would have allowed contact tracing and isolation to flatten the epidemic spread. However, if the number of prevalent cases were high, an epidemic was very likely. This was the conclusion of a study that conducted a phylogenetic analysis of the COVID-19 lineages introduced into the UK in 2020.^22^ This study suggested that high travel volumes and few restrictions on international arrivals prior to lockdown led to the establishment of >1000 identifiable UK transmission lineages, which contributed to accelerated epidemic growth that quickly exceeded public health capacity to contain in late February and early March. Notably, most of these cases were deemed to have come from European countries such as France and Spain.

In developing a comprehensive explanation of East-West differences in excess deaths and life expectancy losses from COVID-19, it is useful to distinguish between proximal and distal influences (Figure 1). Proximal ones are those that influence the spread of the pandemic in each country, such as promulgation of and population compliance with NPIs and vaccine uptake including hesitancy and availability, and prevalence of infection at the time of initial lockdown. Distal influences are features that may differ between Eastern and Western European countries that originate in the different post-war histories of the two blocs which may have influenced how governments and individuals responded to the pandemic. These include differences in public trust in government and science leading to a varying degree of compliance with government advice or regulation aimed at reducing levels of transmission as well as affecting differences in levels of vaccine hesitancy. We discuss each of these in turn below.

The underlying difference in transport connectivity between East and West can be seen as part of the longer-term legacy of the historical divisions across the European continent in the post-war period. This reflects differences in patterns and intensity of international business and commerce as well as favoured tourist destinations. The development of major airline hubs in the UK, France, Germany, Spain, and Italy has occurred over many decades. In Eastern Europe countries, however, the entry and exit of people was severely constrained up until the fall of the Berlin Wall and the collapse of the Soviet Union.

The summer of 2020 was accompanied by a relaxation of many of the most stringent restrictions on movement and contact between people, both within and between European countries, which drove a resurgence of infections in the early autumn.^23^ In particular, it would have resulted in the seeding of new infections in Eastern European countries that had been avoided in the first wave of the pandemic and which eventually drove the substantial peaks of excess mortality in these countries from October 2020 onwards.

In 2021, Eastern European countries lagged behind those of Western Europe in terms of pace and ultimate level of population coverage of COVID-19 vaccination (Supplementary Figure S7). We have found that this is the most influential factor in reducing the size of the East-West difference in life-expectancy losses. Higher levels of vaccination hesitancy in Eastern European countries, particularly in Russia and Bulgaria may be particularly relevant.^24–27^ Steinert et al.^28^ found no adequate response to messages about the benefits of vaccination in several countries including Bulgaria and Poland, which was not the case in Germany and the UK.

There has been much scientific and political interest in quantifying the extent to which NPIs played a role in reducing the impact of the pandemic. ^29–31^ Our analysis found that differences in NPIs as measured by mean levels of the stringency index in each country over 2021 did not explain any of the East-West differences in life expectancy losses in 2021. However, the stringency index we used has many serious shortcomings in this context. Firstly, the relationship between the level of NPIs and SARS-CoV-2 infection was iterative and bidirectional: while the stringency of NPIs would depend on the level of SARS-CoV-2 in the community, the latter would also inform the subsequent level of NPI stringency. Secondly, the stringency index we used summarises the government’s intent, rather than actual uptake and compliance. Thirdly, in retrospect the stringency index was a simple sum of the NPI measures, despite the fact that not all the NPIs have the same impact on the transmission of SARS-CoV-2.

What is clear is that the massive fall in cross-border movement that occurred quickly in Europe and elsewhere from March 2020 had a major impact on the spread of the virus.^29^ This is the necessary corollary of our conclusion that it was the differences in the prevalence of SARS-CoV-2 infection in European countries that are key to understanding the later excess mortality peaks in Eastern compared to Western countries in 2020.

The extent to which the experience of communism in Eastern Europe in the 20^th^ century has had a persistent negative effect on trust at many different levels of society has been extensively studied.^32^ A large multi-regional study conducted in 2020 across 16 countries found that the four countries from Central and Eastern Europe showed the lowest level of trust in government.^33^ In our analysis, trust in government, and to a smaller degree, trust in science, appears to explain a component of East-West differences in life expectancy losses in 2021. This could be through a moderating influence on the willingness of individuals to comply with recommended or required behaviour changes aimed at reducing person-to-person transmission. It could also operate through an association of trust with vaccine uptake.

Differences in the degree to which countries in Eastern and Western Europe tend to enforce laws and regulations in general may have an impact on differences in life expectancy losses.^14^ This may operate both through the extent of policing rule-breaking social gatherings as well as business breaches of regulations. The level of trust in institutions as a factor that could influence the impact of the pandemic on populations has been explored in a number of studies.^33–35^ A study looking at factors influencing adherence to NPIs, and its change over time as populations became fatigued, found that reductions in adherence to physical distancing occurred to a smaller degree in countries with high interpersonal trust.^36^

Pre-pandemic research on vaccine hesitancy has identified trust as an important factor.^37^ This is consistent with the findings of studies of COVID-19 vaccine uptake^38^, although another study specifically of Eastern Europe using a questionable measure of trust was inconclusive.^39^ However, our results suggest that the combination of vaccine coverage together with trust in government reduces by half the observed East-West differences in life expectancy losses, indicating that trust in the government may be important above and beyond its relationship to vaccine coverage, possibly through its influence on, among other factors, compliance with NPIs.

Our analysis has some weaknesses. The analysis of flight connectivity was based on the numbers of flights, rather than actual passenger numbers, which would provide the most appropriate metric for assessing the probability of case import. The lack of data on passengers also meant that we could not differentiate between people who landed and entered the destination population versus those who were in transit and traveled onwards to another country. This latter limitation may explain why Germany is such an outlier in Supplementary Figure S3. Frankfurt is known as having one of the world’s largest transit hubs by passenger volume. Finally, the cross-border spread of infection is a function of land transport and not just flying, although train, bus, and car travel would predominantly be between adjacent countries.

The variables we used as proxies for the factors that might underlie East-West differences in losses in 2021 may only partially capture the underlying concepts. In addition, we do not have proxy measures for some of the key pathways shown in the conceptual diagram, including the availability and effectiveness of treatments for people with COVID-19. Moreover, we did not have measures of individual and institutional behaviour change that occurred in response to government advice/stipulation or simply individual responses to information available through the news and social media about the pandemic. However, we have used more upstream (pre-pandemic) factors, such as general tendency for enforcement as well as trust in government and science, which we have regarded as potentially important general influences on behaviours of individuals and institutions.

Our conceptual model of life expectancy losses in 2021 does not take into account the fact that there are very likely to have been differences between Eastern and Western European countries in the levels of immunity to SARS-CoV-2. Data on population-based seroprevalence of antibodies in Europe at the start of 2021, or at any other point in the pandemic, is very limited.^40^ However it is notable that in England, which had one of the earliest and most intense peaks in COVID-19 excess deaths, the prevalence of immunity in the population in mid-July 2020 was surprisingly low at around 6%.^41^ Other time series data from the UK show that the proportion of the population with levels of immunity only started to increase above this level once mass vaccination began in early 2021.^42^ However, it is notable that in several Western European countries which had late first peaks in autumn 2020 (e.g. Austria, Switzerland, Germany, Denmark), like all of the Eastern countries, nevertheless ended up with far lower excess mortality and life expectancy losses in 2021. Overall, we conclude that low rates of naturally acquired immunity at the start of 2021 would be unlikely to explain much of the East-West difference in life expectancy losses in 2021.

In conclusion, the main early impact of the pandemic in the spring of 2020 was felt in several Western European countries, with none of the former communist countries of Eastern Europe experiencing a major peak in excess mortality until the autumn of 2020. Despite this, the overall level of losses up to the end of 2021 in life expectancy and excess death rates were far higher in the East compared to the West. We argue that both of these distinctive features of the East-West difference can be ultimately related to the differences in the post-war history of the two blocs. Of particular importance is the lower degree of international transport connectivity of Eastern countries which is likely to explain a lower prevalence of infection in March 2020 when lockdowns were initiated around the world. Lower levels of trust in government and science, and willingness to enforce laws and regulations in the East are likely to be behind the far higher excess mortality rates seen in Eastern than in Western countries in 2021. This appears to have been importantly mediated principally through lower levels of vaccine coverage as well as population adherence to NPIs.

## Contributors

V.M.S. and S.T. conceived the study. V.M.S., S.T., and D.J. collected, pre-processed and validated the data with the help of N.M-J who obtained and processed the flight connectivity data. V.M.S. carried out the demographic and statistical analyses with contributions from D.J. and S.T. S.T. and D.A.L. drafted the manuscript. V.M.S., S.T., D.J., N.I., and D.A.L. contributed to subsequent versions, and to the interpretation of the data and results. All authors reviewed and approved the final version of the manuscript.

## Funding

S.T. acknowledges support from the Australian Research Council (DP210100401). N.I. acknowledges support from the UK National Institute for Health and Care Research (HDRUK2022.0313). The funders had no role in study design, data collection and analysis, decision to publish or preparation of the manuscript.

## Competing interests

None declared.

## Patient and public involvement

Patients and/or the public were not involved in the design, conduct, reporting, or dissemination plans of this research.

## Patient consent for publication

Not applicable.

## Ethics approval

Not applicable.

## Provenance and peer review

Not commissioned; externally peer reviewed.

## Data availability statement

The raw data originated from the open access sources listed in the References. We also provide figures and tables data in separate Excel files (https://github.com/VMSdemo/East-West-contrast-in-life-expectancy-losses-in-2020-21). Some of the Excel files contain calculations (including decomposition of life expectancy losses) and final data manipulations.

## Acknowledgements

We would like to thank Professor John Edmunds for his valuable insights and suggestions on an earlier draft of the paper.

## Notes

### Competing Interest Statement

The authors have declared no competing interest.

### Summary of Updates

We have made some important changes in the analytical part of the paper and in the discussion.

