## Supplementary for "East-West mortality disparities during the COVID-19 pandemic widen the historical longevity divide in Europe: an international comparative study" (from another version of this preprint: version 1, 2023-11-09)

### **Supplementary Methods**

**Mortality data**

In calculations of the annual life expectancy in 2000-2021 and life expectancy losses in 2020-21, we used country-specific death counts and population exposures from the HMD(1) core that allowed us to compute sex- and age-specific (0,1-4, 5-9, …, 100+) death rates for every year from 2000 to 2021. For the most recent years for which the final annual data were not yet published for some countries in the HMD core, these death rates were computed from weekly age-specific mortality data from the Input section of the Short-Term Mortality Fluctuations series (STMF).(2)

STMF series was also used as a primary source for assessing weekly series of crude death rates (CDRs) from 2000 to 2021 and excess crude death rates (ECDRs) in 2020-21.

**Calculations from weekly mortality data: the weekly death rates, the timing of the earliest major peaks, and the total excess death rate in 2020-21**

The baseline (expected) CDRs by weeks of 2020-21 were extrapolated from weekly CDRs in 2005-2019 by using a variant of the Serfling model that included a quadratic annual trend and two sin-cosine harmonics for seasonal fluctuations.(3)^,^(4)

${CDR}_{y,i}=a+by+dy^{2}+s_{1}\sin\left( \frac{2\pi i}{52} \right)+c_{1}\cos\left( \frac{2\pi i}{52} \right)+s_{2}\sin\left( \frac{2\pi i}{26} \right)+c_{2}\cos\left( \frac{2\pi i}{26} \right)+\varepsilon_{y,i}$,	(1)
with calendar year *y*, week *i*, parameters *a*, *b*, *s*_1_, *c*_1_, *s*_2_, *c*_2_, and error term epsilon.

For each country, we defined the earliest major peak of excess mortality as the first local maximum of weekly ECDRs exceeding the 95% percentile of weekly ECDRs over 2020-2021. In Russia and Estonia, we defined the earliest major peak slightly differently. Due to the very large size of Russia, the timing of the pandemic differed between regions of the country. Consequently, the distribution of weekly ECDRs was considerably more dispersed than such distributions in other countries. To address this peculiarity, we used 90% threshold instead of 95%. In Estonia, the first ECDR peak exceeding the 95% threshold took place on the 25th week (June, 21-27) of 2021. However, this peak was caused by a heat wave.(5) Therefore, we took the next mortality maximum (week 44) as the first major ECDR peak in Estonia.

The total ECDR values for the groups East and West were computed as averages of country-specific life expectancy losses.

We created a geographical map of the peaks that illustrates the spread of excess mortality across European countries. Another map showed the distribution of the total ECDR of 2020-21 across Europe. The maps were built in ArcGIS 10.8.2.

**Calculations from annual mortality data: the life expectancy losses in 2020-21 and their decomposition by broad age group**

Abridged life tables for all 28 populations for years from 2000 to 2021 were estimated from the observed age-specific death rates by applying the standard life table technique.(6) The life expectancy at birth series was extracted from these life tables.

The Lee-Carter model was applied for forecasting the age- and sex-specific death rates and abridged life tables for 2020 and 2021 using the 2005-2019 as the reference (fitting) period. The choice of the period was based on the previously conducted sensitivity analysis.(2, 7) This analysis demonstrated that short reference periods such as 2015-19 and (to a lesser extent) 2010-19 may lead in some populations to poorer fit of the Lee-Carter model and may also result in underestimation of life expectancy losses.

We then extracted from the latter life tables the baseline life expectancies at ages 0 and 65, interval life expectancy between exact ages 0 and 65, and the probability of survival to age 65. We also estimated 95% confidence limits for these quantities by carrying out 2000 bootstrap simulations nested on prediction errors of the age-specific death rates.

The life expectancy losses were computed as the differences between the baseline and the observed life expectancy values in 2020 and 2021.

To quantify the contribution of ages 0-64 to the overall life expectancy loss using *l*_x_ and *e*_x_ functions at ages 0 and 65 only, we applied the following formula:

$\delta_{0-64}= \left[ \left( l_{0}^{*}e_{0}^{*}-l_{65}^{*}e_{65}^{*} \right)-\left( l_{0}e_{0}-l_{65}e_{65} \right) \right]+\frac{1}{2}\cdot\left[ \left( l_{65}^{*}-l_{65} \right)e_{65}^{*}-\left( l_{65}-l_{65}^{*} \right)e_{65} \right]$, (2)

where $e_{0}$/$e_{0}^{*}$and $e_{65}/e_{65}^{*}$ are observed/baseline life expectancies at age 0 and age 65 in the year 2020 or 2021; $l_{0}$/$l_{0}^{*} \mathrm{and}l_{65}/l_{65}^{*}$- observed/baseline life table numbers of survivors to ages 0 and 65.

The two additive terms on the right-hand side of Eq. 2 are equal to 1) the lifetime loss between exact ages 0 and 65 produced by mortality differences at ages 0 to 64; 2) the lifetime lost at ages higher or equal to 65 produced by mortality differences at ages 0 to 64.

The contribution of ages 65+ is estimated as the difference between the overall life expectancy loss$\delta$ and the one imposed by younger ages $\delta_{0-64}$:

$$\delta_{65+}=\delta-\delta_{0-64}$$

**Computation of contributions of relative mortality excess and baseline mortality to the total East-West differences in the life expectancy losses**

The life expectancy losses in a *target* Eastern population and a *reference* Western population are

$\delta_{E}= {e_{0, E}^{*}-e_{0,E}=e}_{0}\left( \boldsymbol{M}_{E}^{*} \right)-e_{0}\left( \boldsymbol{M}_{E} \right) ,$	(3a)

$\delta_{W}= e_{0, W}^{*}-e_{0,W}={e_{0}\left( \boldsymbol{M}_{W}^{*} \right)-e}_{0}\left( \boldsymbol{M}_{W} \right)$,	(3b)

where ${\boldsymbol{M}_{E}\boldsymbol{,}\boldsymbol{M}_{W,}\boldsymbol{M}}_{E}^{*}, \boldsymbol{M}_{W}^{*}$are the observed and the baseline vectors of age-specific death rates, and the life expectancy at birth is expressed as a scalar function of these vectors.

The East-West difference in the life expectancy losses ${\Delta_{E-W}=\delta}_{E}-\delta_{W}$ is considered as being dependent on: A) a larger relative mortality excess due to a larger mortality increase in the East compared to the West (*Change* factor); B) a higher level of baseline death rates in the East compared to the West (*Level* factor). The East-West difference $\Delta_{E-W}$ can be presented as a sum of the Change and Level components

${\Delta_{E-W}=\Delta}_{C}+\Delta_{L}$.	(4)

The calculation formulae for the components $\Delta_{C}$ and $\Delta_{L}$ is given in Shkolnikov et al.(8)

**Estimation of associations between the East-West differences in life expectancy losses with explanatory variables across countries in 2021**

The set of explanatory variables included proxy variables for the factors shown in the conceptual diagram (Fig. 7):

- Percentage of fully vaccinated as of the 1^st^ September of 2021(9);
- Mean Stringency Index over weeks of 2021(10);
- Percentage of people who trust the national government reported in 2020(11);
- Percentage of people who trust science reported in 2020(11);
- Regulatory Enforcement Index in 2021.(12)

To check the applicability of linear OLS models, we checked the normality of variables’ distributions by visual examinations and carried out statistical tests for linearity and heteroscedasticity (Breusch-Pagan/Cook-Weisberg test) of relationships between the life expectancy losses and each explanatory variable.

To obtain normality of the vaccination variables we trimmed one outlying point for Bulgaria by replacing the original value with the average of the 1st and the 5th percentiles of the distribution of this variable. We have not found any evidence for the non-normality of the residuals. There was also no evidence of non-linearity of the relationships. Finally, we found evidence of heteroscedasticity in the relationships between the male and female life expectancy losses on one side and the regulation enforcement and trust in the national government variables on the other. We addressed it by using robust confidence limits in the corresponding regression models.

After completing the tests, we regressed (using robust standard errors) male and female life expectancy losses on vaccination as of the 1^st^ of December 2021, trust in government, trust in science, and regulatory enforcement index. Finally, we examined the model East-West difference in male and female life expectancy losses (presented by the East-West dummy) and how this difference attenuates in response to adjustment for single explanatory variables and for two-variable combinations. The use of the two-variable models was justified through the fit criteria: the root mean squared error (RMSE), and the Bayesian information criterion (BIC).

### **Supplementary Figures**

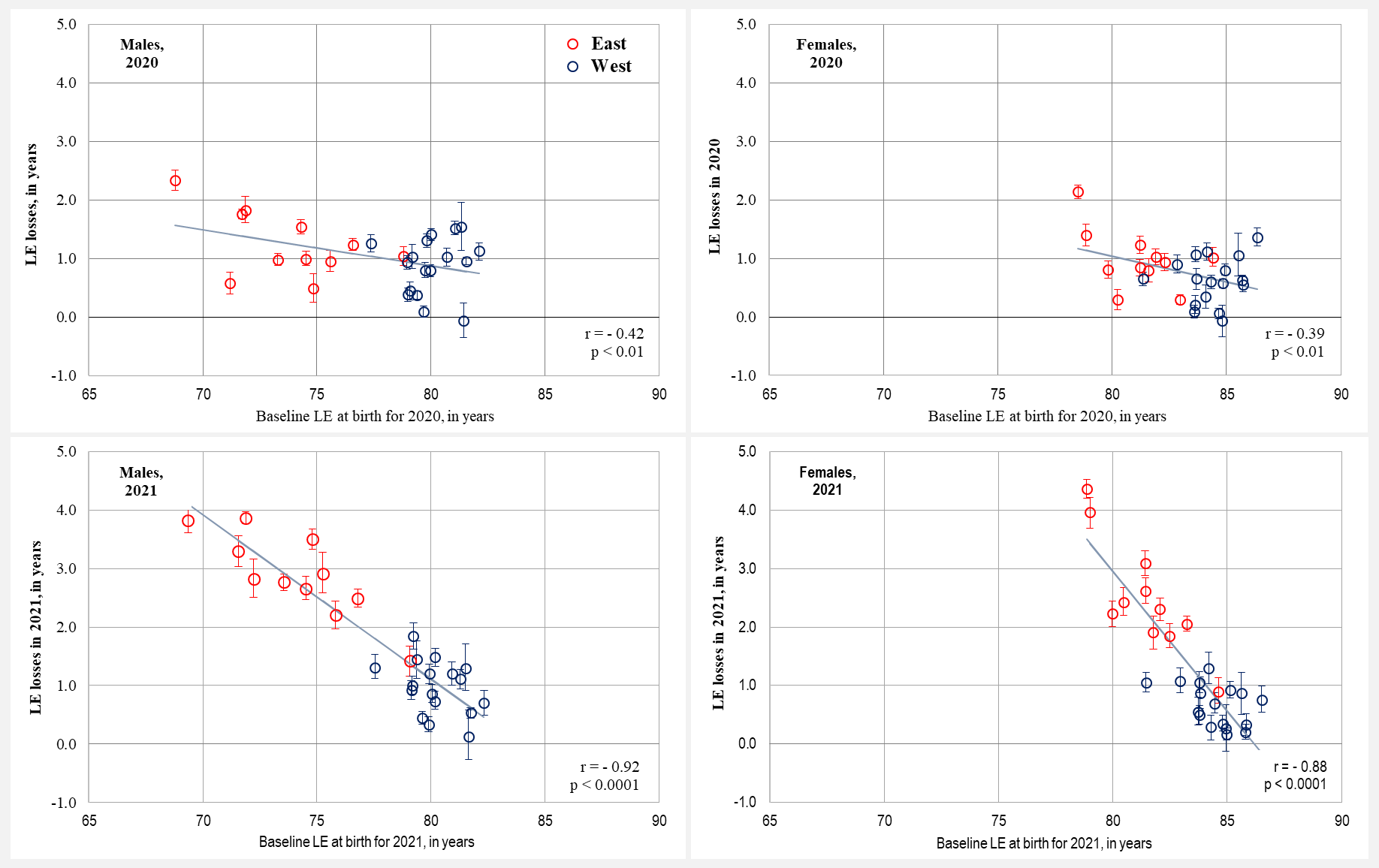

**Supplementary figure S1. Associations between life expectancy losses (95% CI) and predicted life expectancies, by year and sex.**

Life expectancy loss shows the difference between observed and expected (in the absence of pandemic) life expectancies. A pronounced East-West difference in life expectancy losses became apparent in 2021. Lower level of life expectancy was strongly associated with higher life expectancy loss.

Data shown in this Figure is provided at https://github.com/VMSdemo/East-West-contrast-in-life-expectancy-losses-in-2020-21

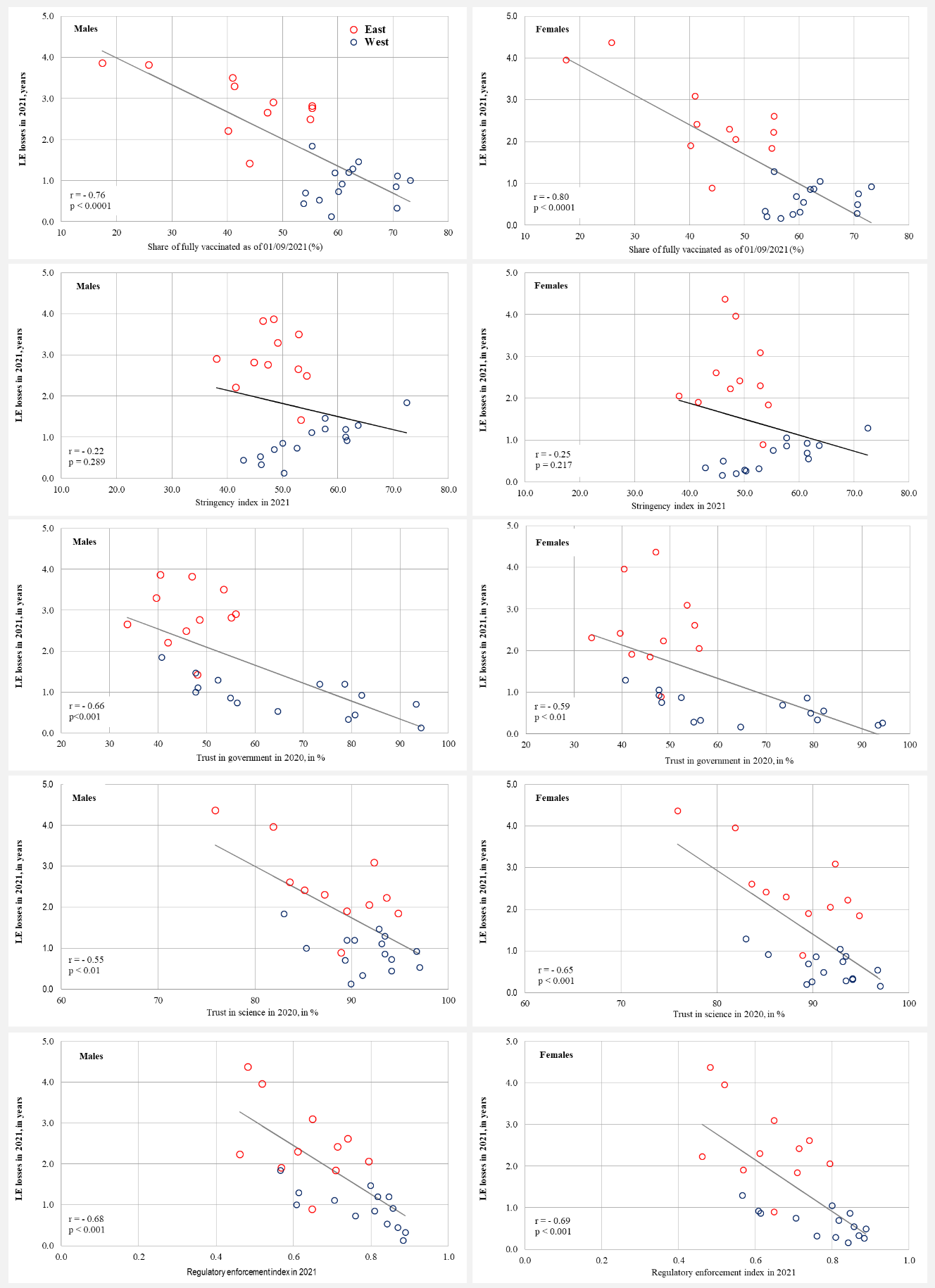

#### **Supplementary figure S2. Associations between sex-specific life expectancy losses in 2021 with single explanatory factors across countries.**

OLS regression lines and Pearson correlation coefficients are shown in the panels. Life expectancy losses are negatively correlated across countries with vaccination, trust, and regulatory enforcement variables. No statistically significant correlations found for the stringency index.

Data shown in this Figure is provided at https://github.com/VMSdemo/East-West-contrast-in-life-expectancy-losses-in-2020-21

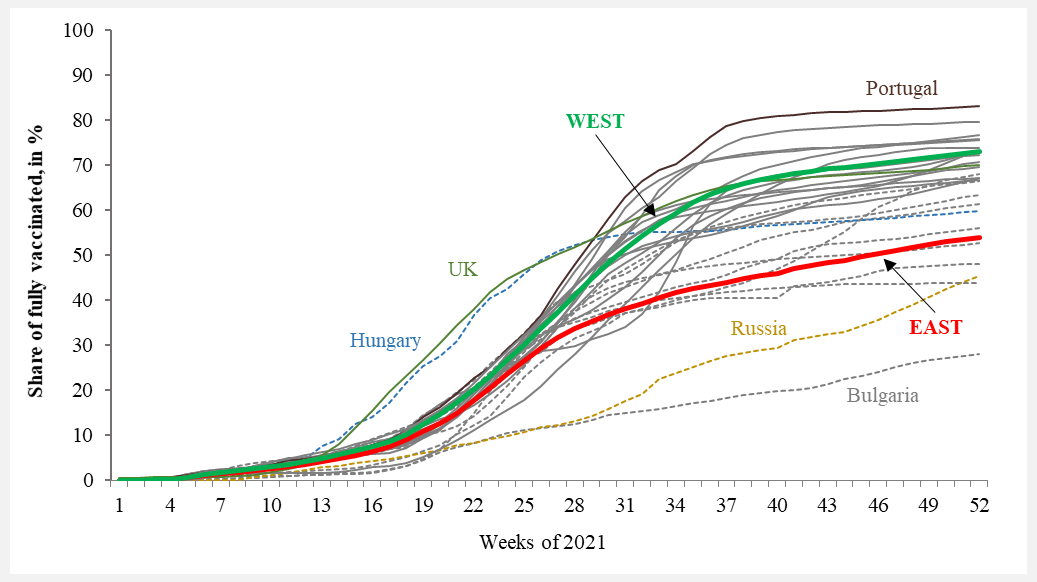

#### **Supplementary figure S3. Cumulative prevalence of full vaccination by weeks of 2021 across countries and country groups.**

Lower vaccination in Eastern European countries particularly in Bulgaria and Russia. A remarkable exception for higher vaccination prevalence in Hungary.

Data shown in this Figure is provided at https://github.com/VMSdemo/East-West-contrast-in-life-expectancy-losses-in-2020-21

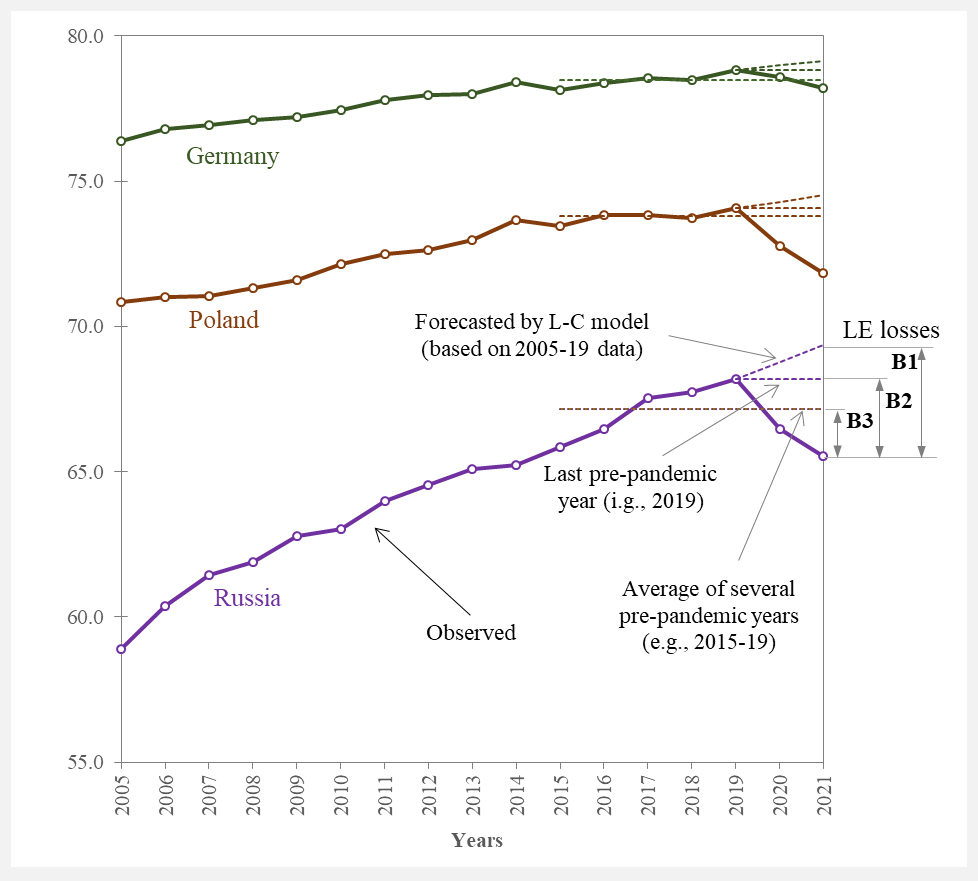

#### **Supplementary figure S4. A graphic presentation of different approaches towards estimating baseline mortality and corresponding life expectancy losses in 2020-2021: an example of Russia, Poland, and Germany.**

B1: Baseline 1 - Lee-Carter predicted life expectancy from age-specific death rates in 2005-2019. B2: Baseline 2 - life expectancy in 2019; B3: Baseline 3 - average life expectancy in 2015-19. Use of Baselines 2 and 3 leads in underestimation of life expectancy losses if life expectancy increases with time.

Data shown in this Figure is provided at https://github.com/VMSdemo/East-West-contrast-in-life-expectancy-losses-in-2020-21

### **Supplementary Tables**

#### **Supplementary table S1. Populations and regional groups under study**

| Eastern Europe (East),  11 countries | | Western Europe (West),  17 countries | |
| --- | --- | --- | --- |
| Country name | **Abbreviation** | **Country name** | **Abbreviation** |
| Bulgaria | BGR | Austria | AUT |
| Czechia | CZE | Belgium | BEL |
| Estonia | EST | Switzerland | CHE |
| Croatia | HRV | Germany | DEUTNP |
| Hungary | HUN | Denmark | DNK |
| Lithuania | LTU | Spain | ESP |
| Latvia | LVA | Finland | FIN |
| Poland | POL | France | FRATNP |
| Russia | RUS | England and Wales | GBRTENW |
| Slovakia | SVK | Northern Ireland | GBR_NIR |
| Slovenia | SVN | Scotland | GBR_SCO |
|  |  | Greece | GRC |
|  |  | Italy | ITA |
|  |  | Netherlands | NLD |
|  |  | Norway | NOR |
|  |  | Portugal | PRT |
|  |  | Sweden | SWE |

#### **Supplementary table S2. Contribution of relative mortality excess and baseline mortality to the East-West difference in life expectancy losses in 2021**

|  | Males | Females |
| --- | --- | --- |
| LE losses in the East (Δ_E_) | 3.02 | 2.64 |
| LE losses observed West (Δ_W_) | 0.99 | 0.67 |
| Differences in LE losses between East and West (Δ_E-W_) | 2.03 | 1.97 |
| LE losses due to larger relative mortality excess (Δ_C_) | 1.78 | 1.86 |
| LE losses due to higher baseline mortality (Δ_L_) | 0.25 | 0.11 |
| Contribution of the relative mortality excess, % | 87.7 | 94.3 |

LE= life expectancy.

Noteworthy, life expectancy losses for East and West in this table are calculated from life tables with the average age-specific death rates in them. This makes a difference from Table 1 showing means of country-specific life expectancies. In Table S2, these losses are marginally higher by about 0.1 year compared to Table 1.

Data shown in this Table is provided at https://github.com/VMSdemo/East-West-contrast-in-life-expectancy-losses-in-2020-21
